# Resource Allocation for Different Types of Vaccines against COVID-19: Tradeoffs and Synergies between Efficacy and Reach

**DOI:** 10.1101/2021.01.28.21250713

**Authors:** Daniel Kim, Pelin Pekgün, İnci Yildirim, Pınar Keskinocak

## Abstract

**Objective:** Vaccine shortage and supply-chain challenges have caused limited access by many resource-limited countries during the COVID-19 pandemic. One of the primary decisions for a vaccine-ordering decision-maker is how to allocate the limited resources between different types of vaccines effectively. We studied the tradeoff between efficacy and reach of the two vaccine types that become available at different times.

**Methods:** We extended a Susceptible-Infected-Recovered-Deceased (SIR-D) model with vaccination, ran extensive simulations with different settings, and compared the level of *infection attack rate* (IAR) under different reach ratios between two vaccine types under different resource allocation decisions.

**Results:** We found that when there were limited resources, allocating resources to a vaccine with high efficacy that became available earlier than a vaccine with lower efficacy did not always lead to a lower IAR, particularly if the former could vaccinate less than 42.5% of the population (with the selected study parameters) who could have received the latter. Sensitivity analyses showed that this result stayed robust under different study parameters.

**Conclusions:** Our results showed that a vaccine with lower resource requirements (wider reach) can significantly contribute to reducing IAR, even if it becomes available later in the pandemic, compared to a higher efficacy vaccine that becomes available earlier but requires more resources. Limited resource in vaccine distribution is significant challenge in many parts of the world that needs to be addressed to improve the global access to life-saving vaccines. Understanding the tradeoffs between efficacy and reach is critical for resource allocation decisions between different vaccine types for improving health outcomes.

## INTRODUCTION

In December 2019, the novel coronavirus (SARS-CoV-2), which causes coronavirus disease 2019 (COVID-19), was first detected in Wuhan, China. As of October 2021, approximately 236 million COVID-19 cases have been reported worldwide [1]. Despite the development of effective vaccines at unprecedented speed and high vaccination rates in some countries, vaccine availability remains scarce and vaccination rates remain low in many countries; for example, only approximately 2.3% of people in low-income countries received at least one dose of vaccine as of October 2021 [2].

The procurement and dissemination of vaccines, especially the mRNA vaccines, which require ultra-cold storage, have been particularly challenging in low-income countries [3, 4]. Even before the vaccines were produced, high-income countries had purchased or reserved large amounts of vaccines [5]. Consequently, low- and middle-income countries had difficulty in procuring vaccines early and faced varying prices and unstable supply chains, similar to what was also experienced during the 2009 H1N1 pandemic [6, 7]. Key ingredients, such as lipid nanoparticles used in the production of high-efficacy mRNA vaccines have been in short supply [8, 9]. The limited availability of cold-chain storage and logistics capacity can impede the distribution of mRNA vaccines in many regions. By contrasts, non-mRNA vaccines, such as adenovirus-vectored vaccines, can be transported and stored at lower temperatures, enabling easier distribution and broader reach with less resources.

In general, the procurement and distribution of mRNA versus non-mRNA vaccines require more “resources” such as financial resources, logistics and storage capacities, or healthcare facilities and personnel. Limited vaccine supply in resource limited settings and estimated vaccine shortage for 2022 are real daily life challenges that we have to resolve to improve the global access to life-saving pandemic vaccine [10, 11]. Hence, there is often a tradeoff between efficacy and reach across different vaccines because the resource requirements impact the speed and reach of vaccine distribution efforts, and, consequently, impact how many people gain timely access to vaccination and the level of protection in the population. A recent modeling study considered various efficacies for a single type of vaccine [12, 13], and showed that the wider reach (i.e., lower resource requirements) of the vaccine could result in significant reductions in total infections.

The main goal of this study is to understand this tradeoff between efficacy and reach while developing and deploying vaccine procurement and distribution plans, i.e., how to allocate limited resources between two types of vaccines: (i) a high efficacy vaccine that becomes available earlier but requires more resources, versus (ii) a low-efficacy vaccine that becomes available later but requires less resources. We developed an extended Susceptible-Infected-Recovered-Deceased (SIR-D) simulation model to analyze and assess the impact of resource allocation decisions across two types of vaccines on population health outcomes, such as the proportion of the population infected during the course of the disease. This study provides insights to decision-makers regarding resource planning and allocation when multiple types of vaccines are available during a pandemic. The results suggest that prioritizing the high-efficacy vaccine in resource allocation does not always lead to the best health outcomes; under resource constraints, utilizing a combination of high- and low-efficacy vaccines might reduce the percentage of the population infected and reduce the infection peak.

## METHODS

### Two Types of Vaccines

We considered two types of single-dose vaccines that become available at different times. The vaccine with high efficacy (vaccine-H) becomes available sooner than the vaccine with lower efficacy (vaccine-L). A resource of *K* units is available daily (*K*=1 million in the simulation), and a fixed proportion *a*_*H*_, *a*_*L*_ ∈ [0,1] of the capacity *K* is allocated to vaccine-H and vaccine-L, respectively, where *a*_*H*_ + *a*_*L*_ = 1. For a given *K, K* people can receive vaccine-L (i.e., one unit of resource is needed for one person to receive vaccine-L), whereas only a *λK* people (*λ* < 1) can receive vaccine-H. We denote *λ* as the *reach ratio*, where lower *λ* values indicate higher resource requirements for vaccine-H relative to vaccine-L. Hence, given daily capacity *K*, reach ratio *λ*, and allocation decisions *a*_*H*_, *a*_*L*_ *(a*_*H*_ + *a*_*L*_ = 1), the number of people who can receive vaccine-H and vaccine-L daily are *a*_*H*_ *λK* and *a*_*L*_*K*, respectively.

In the main scenario, we set the efficacy of vaccine-H at 90% and vaccine-L at 70%, and vaccine-H becomes available three weeks earlier than vaccine-L. We assumed that during the period when only one vaccine type is available, any unused daily resources are lost.

### Compartmental epidemiological model

In the extended SIRD model, each person is in one of the following compartments at a given time: *Susceptible* (*S*), *Susceptible-i* (*S*_*i*_, *i* = H or L), *Symptomatic-Infected* (*I*^*S*^), *Asymptomatic-Infected* (*I*^*A*^), *Quarantined* (*Q*), *Vaccinated*-*i* (*V*_*i*_), *Recovered* (*R*), and *Deceased* (*D*). *Susceptible* (unvaccinated) population transitions to one of the *Symptomatic-Infected* (*I*^*S*^) or *Asymptomatic-Infected* (*I*^*A*^) compartments after *infectious contact* with either infected population. Depending on the resource allocation decisions and vaccine efficacy, a proportion of Susceptible population who receives vaccine-*i* (*i* = H or L) transitions to *Vaccinated*-*i* (*V*_*i*_), whereas the others transition to *Susceptible*-*i* (*S*_*i*_; those who received vaccine-*i* but did not develop immunity). Symptomatic-Infected population transitions to one of the *Quarantined* (*Q*), *Recovered* (*R*) or the *Deceased* (*D*) compartments. Asymptomatic-Infected population transitions to *Recovered* (*R*) compartment. The transition diagram of the extended SIRD model is depicted in Figure 1.

**Figure 1:**
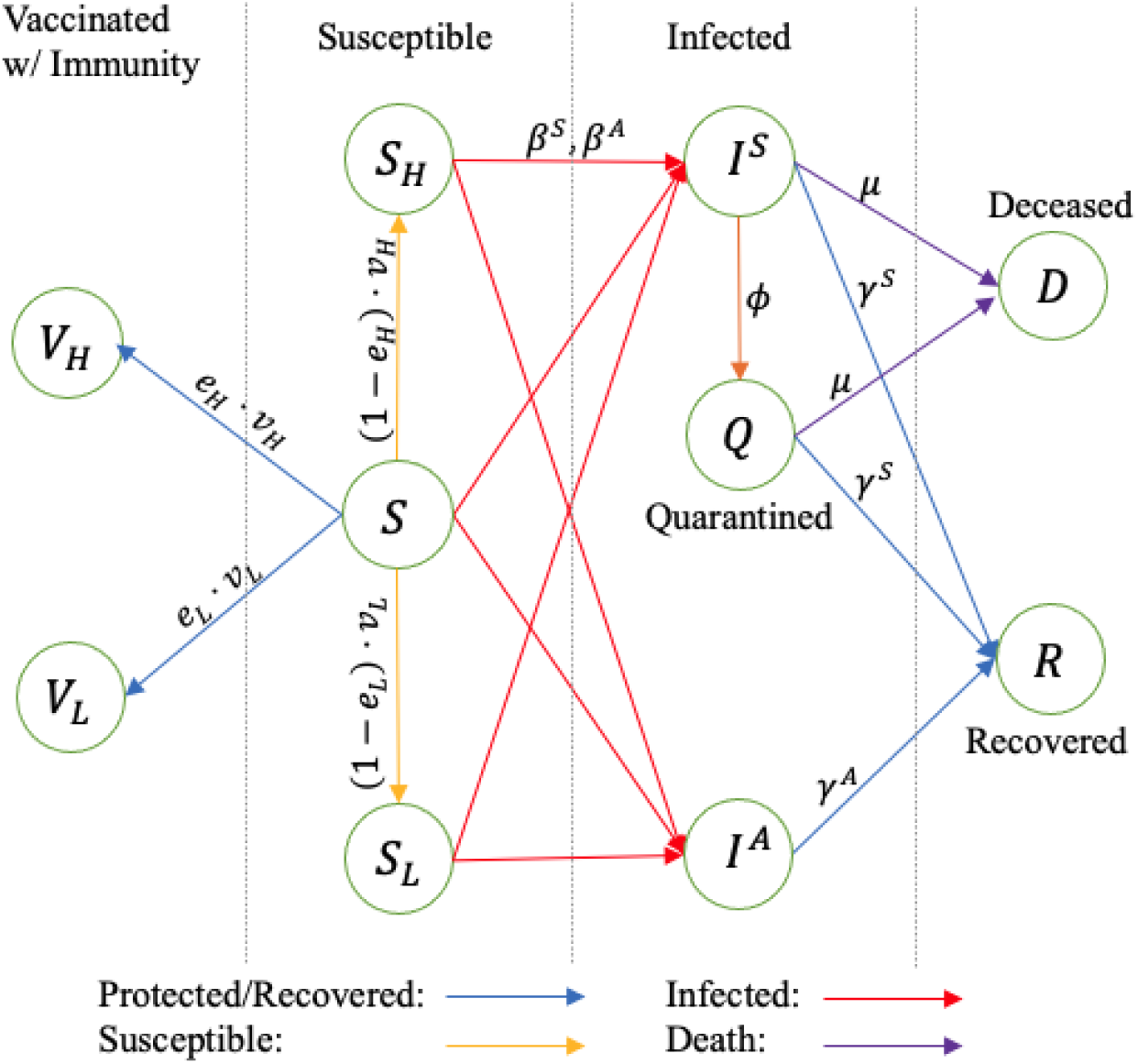
Transmission diagram of the extended SIR-D model, in which the population is stratified based upon the epidemiological status; : Transmission rate due to infectious contacts between a susceptible individual and either a symptomatic or asymptomatic patient; : Efficacy of vaccine-H and vaccine-L; : ratio of the daily vaccine capacity to the size of the susceptible population (i.e., and); : Self-isolation rate; : Recovery rate of a symptomatic or asymptomatic patient; : Death rate of a symptomatic patient.

The parameter values in the SIR-D model were chosen based on the SARS-CoV-2 characteristics. The Centers for Disease and Prevention (CDC) in the United States estimated that 70% of COVID-19 infections are symptomatic ([14]. The durations of the mean presymptomatic infectious period, the median asymptomatic infectious period, and the mean time from symptom onset to two negative RT-PCR tests are estimated as 6 days [15], 6.5–9.5 days, and 13.4 days [16], respectively. Hence, we set the recovery rates of asymptomatic patients () and symptomatic patients () at 1/8 and 1/16, respectively. The CDC reported that the number of days from symptom onset to SARS-CoV-2 test ranges between 0 and 4 days [14]. Considering the time until the test result becomes available and the number of people getting tested, we set the quarantine rate (*ϕ* = 1/12), by which the symptomatic infectious population (*I*^*S*^) move to the Quarantined compartment (*Q*). Infection fatality rate (IFR-S) for symptomatic infectious population is estimated as 1.3% in the United States [13] and lower in a typical low-income country with younger population [17, 18]. Hence, we set the death rate (*µ*) to be 0.0015, at which IFR-S is approximately 1.07% in the simulations without the vaccines. The infectivity of a disease, represented by reproduction number (*R*_0_), is estimated as 2.5 by [14]; the symptomatic-transmission rate (*β* ^*S*^) is set at 0.21 in the main scenario of the simulation, and the asymptomatic-transmission rate (*β* ^*A*^) is set at 75% of the symptomatic-transmission rate [19].

We ran the simulation using R-software with a population size of 330 million. Since our main goal is to analyze the impact of resource allocation of multiple types of vaccines, we started the simulation only after vaccine-H became available (Day 1). For the initial population size (immediately before vaccine-H becomes available) in each compartment, we set *S* = 94.86%, *I*^*S*^ = 1.02%, *I*^*A*^ = 0.58%, *Q* = 0.34%, *R* = 3.10%, and *D* = 0.01%, motivated by the COVID-19 statistics recorded on December 14, 2020, the first day of the vaccine distribution in the United States [20, 21]. The simulation was run over a one-year planning horizon with *a*_*H*_ = 0 to 1 (*a*_*H*_ + *a*_*L*_ = 1; increments of 0.1), and *λ* = 0.005 to 0.995 (increments of 0.005).

We compared *infection attack rate* (IAR) as the main health outcome, peak day (the day when the peak infections occur), and peak percentage (percentage of the population that is newly infected on the peak day) under different scenarios to evaluate the impact of the resource allocation decisions.

In addition to the main scenario, we performed extensive sensitivity analyses. We simulated 21 additional scenarios with different 1) infectivity of the disease with reproduction numbers (*R*_0_ = 2, 2.25, 2.75 and 3) and corresponding transmission rates, 2) timing when vaccine-L becomes available within the range of 0 to 8 weeks after vaccine-H becomes available (increments of 1 week), and 3) efficacy levels of vaccine-H within the range of 85% to 95% (increments of 1%). We also assessed six alternative scenarios, in which 1) vaccine-H becomes available within the range of 1 to 4 weeks after vaccine-L becomes available (increments of 1 week; four scenarios), and 2) the initial population size of each compartment is different from the main scenario (two scenarios).

## RESULTS

In the main scenario, in the absence of vaccines, approximately 50.18% of the population is infected, the peak day is 39 (from the start of the vaccination), and the peak percentage is 0.65%.

### Infection Attack Rate

Table 1 and Figure 2 show the IAR under different reach ratios and resource allocation decisions. When the reach ratio is low, i.e., *λ* ≤ 0.425: (i) Allocating all resources to vaccine-L (i.e., *a*_*H*_ = 0, *a*_*L*_ = 1) minimizes the IAR. (ii) Comparing the optimal allocation *a*_*L*_ = 1 to allocating all resources to vaccine-H (*a*_*H*_ = 1), the difference between the IARs under *a*_*L*_ = 1 and *a*_*H*_ = 1 increases as *λ* decreases. For example, when *λ* = 0.2 and *λ* = 0.425, the differences in IAR are 4.83% and 0.293%, respectively, corresponding to approximately 16 million infections (Figure S1) and 0.97 million infections that could have been averted by allocating all the resources to vaccine-L versus vaccine-H.

**Table 1:** Infection attack rate (IAR) in percentage under different resource allocation decisions and reach ratios

| $(a_H, a_L)$ | Reach ratio ($\lambda$) | | | | | | |
| --- | --- | --- | --- | --- | --- | --- | --- |
|  | 0.425 | 0.43 | 0.435 | 0.44 | 0.445 | 0.45 | 0.455 |
| (1, 0) | 41.009 | 40.916 | 40.823 | 40.730 | 40.638 | 40.546 | 40.454 |
| (0.9, 0.1) | 40.951 | 40.868 | 40.786 | 40.704 | 40.622 | 40.539 | 40.458 |
| (0.8, 0.2) | 40.901 | 40.828 | 40.756 | 40.684 | 40.612 | 40.540 | 40.468 |
| (0.7, 0.3) | 40.887 | 40.795 | 40.732 | 40.670 | 40.608 | 40.546 | 40.484 |
| (0.6, 0.4) | 40.820 | 40.767 | 40.714 | 40.662 | 40.609 | 40.556 | 40.504 |
| (0.5, 0.5) | 40.789 | 40.746 | 40.702 | 40.659 | 40.615 | 40.572 | 40.529 |
| (0.4, 0.6) | 40.764 | 40.730 | 40.795 | 40.661 | 40.626 | 40.592 | 40.558 |
| (0.3, 0.7) | 40.745 | 40.719 | 40.693 | 40.668 | 40.642 | 40.617 | 40.591 |
| (0.2, 0.8) | 40.730 | 40.713 | 40.796 | 40.679 | 40.663 | 40.646 | 40.629 |
| (0.1, 0.9) | 40.720 | 40.712 | 40.704 | 40.795 | 40.687 | 40.679 | 40.670 |
| (0, 1) | 40.716 | 40.716 | 40.716 | 40.716 | 40.716 | 40.716 | 40.716 |

**Figure 2:**
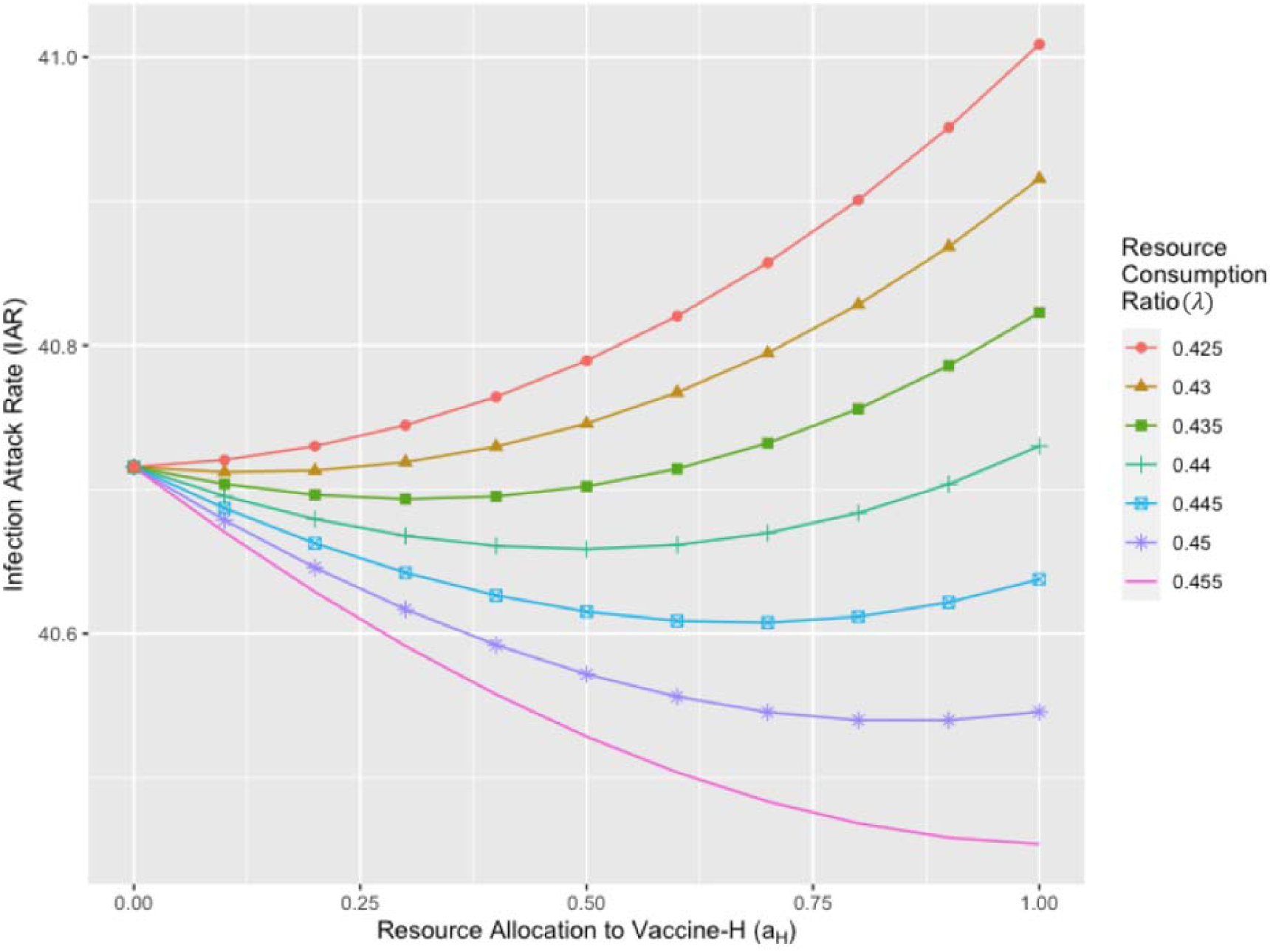
Infection attack rate (IAR) under different resource allocation decisions (with different reach ratios and from top to bottom).

When *λ* ≥ 0.455, allocating the resources entirely to vaccine-H (i.e., *a*_*H*_ = 1, *a*_*L*_ = 0) minimizes the IAR. When the reach ratio falls within 0.425 ≤ *λ* ≤ 0.455, the resources are allocated between vaccine-L and vaccine-H, with the allocation to vaccine-H increasing in *λ*. Figure 3 presents contour plots of IAR under various *λ* and *a*_*H*_ values.

**Figure 3:**
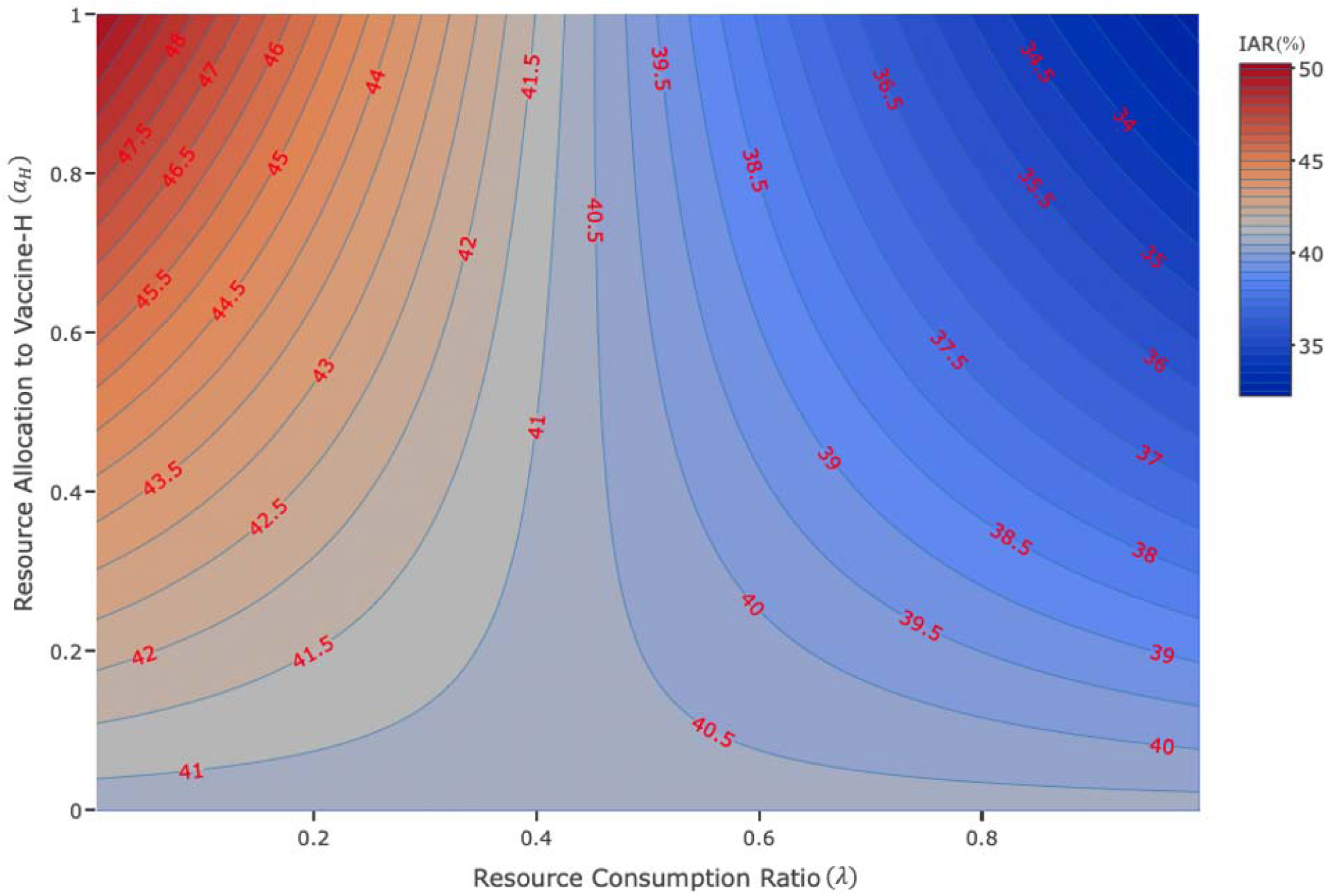
Contour plot of infection attack rate under different reach ratios () and resource allocation decisions ().

### Peak Percentage

Table 12 shows the peak percentage under different reach ratios and resource allocation decisions. The peak percentage is minimized by allocating all resources to vaccine-L and vaccine-H when *λ* ≤ 0.165 and *λ* ≥ 0.21, respectively; when 0.165 ≤ *λ* ≤ 0.21, peak percentage is minimized by splitting the resources between the two types of vaccines.

### Alternative Scenarios

The simulation results of the alternative scenarios with different vaccine efficacies, timings and infectivity of the disease are reported in Supplementary Materials. We observed a similar pattern as in the main scenario in all alternative scenarios. In addition, compared to the main scenario, the maximum below which allocating all resources to vaccine-L minimizes the IAR is lower when the efficacy of vaccine-H is higher, the timing of vaccine-L’s availability is delayed, or the infectivity of the disease is higher. In contrast to the main scenario, if vaccine-H becomes available *later* than vaccine-L, specifically by more than a week, allocating all resources to vaccine-L minimizes the IAR for all.

## DISCUSSION

In this study, we developed an extended SIR-D simulation model and examined the impact of resource allocation decisions across two types of vaccines, namely, a high efficacy vaccine (vaccine-H) that becomes available earlier during the pandemic but requires more resources, and a lower efficacy vaccine (vaccine-L) that becomes available later and requires less resources (i.e., has wider *reach*). For each unit of resource, one person can be vaccinated with vaccine-L whereas *λ* ≤ 1 person can be vaccinated with vaccine-H, where *λ* is defined as the reach ratio. The higher the reach ratio, the wider the distribution of vaccine-H relative to vaccine-L given limited available resources. Our results show that the allocation of limited resources across two vaccine types depends heavily on both the vaccine efficacies and the reach ratio; in particular, there are many scenarios where allocating part or all of the resources to the low efficacy vaccine might lead to better outcomes as measured by the infection attack rate or the peak percentage.

The resource allocation decision is complex due to the tradeoff between efficacy and reach. The more the resources allocated to vaccine-H, the lower the resources remaining for vaccine-L and the lower the total number of people vaccinated (since vaccine-L requires less resources to vaccinate each person). Thus, the reach of vaccine-H needs to be above a certain threshold for it to receive some of the resources.

Our results identified a clear threshold of the reach ratio below which allocating resources entirely to vaccine-L (i.e., *a*_*L*_ = 1) minimizes the IAR. With the selected parameters in the main scenario, the threshold was *λ* = 0.425, indicating that if vaccine-H reaches 42.5% or less of the population who could have received vaccine-L, then vaccine-H is *highly* resource-intensive (Table 1 and Figure 2). When vaccine-H receives some resource allocation, due to its earlier availability and higher efficacy, more people can get vaccinated and build full immunity in the earlier stages of the disease transmission.

However, due to its lower reach level, the rate of decrease in the number of daily infections over time is lower than the rate when the resources are allocated entirely to vaccine-L, which leads to an overall higher IAR in the long term.

While vaccine-L is preferred over a highly resource-intensive vaccine-H, allocating the resources to vaccine-H may achieve a lower IAR when the infectivity of the disease is more severe. Non-pharmaceutical interventions, such as social distancing, mask usage, and school closure, have been deployed during a pandemic and found to be effective in reducing the reproduction number [22-27]. Depending on the various interventions and the disease’s unique infectivity level, different resource allocation decisions need to be made. Our results showed that when the infectivity was high, the implementation of a prompt intervention with vaccine-H was required to prevent a large-scale infection, and, therefore, the threshold of the reach ratio was lower than that in the main scenario (Table S2). This result is consistent with the findings of [24], where the authors studied the impact of the implementation of both non-pharmaceutical interventions and a *single* vaccine type with varying efficacy and coverage.

The timing of vaccine availabilities also influences the threshold of the reach ratio and subsequently the resource allocation decisions. When the timing of vaccine-L’s availability got delayed, we observed that even when it became available after the peak day, the threshold of the reach ratio was lower than that in the main scenario (Table S3 and Figure S2). This indicates that vaccine-H should receive the entire resources since wider infection control with vaccine-L becomes difficult as many get infected until it becomes available. In contrast, when vaccine-H became available *later* than vaccine-L, by more than a week, vaccine-H should receive some resource allocation only when the reach ratio is *λ* ≥ 1, requiring the reach of vaccine-H at least as wide as that of vaccine-L (Table S5). Duijzer et al. (2018) performed an analytical analysis of the resource allocation to two types of vaccines with different efficacies and time of availabilities using a simple SIR model [28]. Under the setting in which vaccine-H became available later than vaccine-L and both vaccines had the same *reach* levels, they showed that allocating the resources entirely to the vaccine with the earlier availability was favored if the later vaccine became available too late, which is consistent with the results of our alternative scenarios.

In addition, we observed a lower threshold for the reach ratio when the availabilities of both vaccines got delayed. Many low- and middle-income countries receive their first batches of vaccines much later – after the disease has already spread widely. During this delay-period, the active and cumulative numbers of infection increase, putting the more susceptible population at risk of infection. Consequently, allocating more resources to the resource-intensive vaccine-H, which becomes available early, may bring larger health benefits than vaccine-L (Table S6 and S7). Thus, despite the difference in the efficacy levels of each vaccine type, depending on the timing of availability and the reach ratio, the decision of which vaccine type should be allocated more resources to minimize the IAR changes significantly. Policymakers must consider the timing of availability for each vaccine type and see if the reach of the later vaccine is wide enough to slow down the infections that are expected to occur during the delay-period.

While the effort of developing a higher efficacy vaccine is significant, our study showed that the impact of increasing efficacy level for vaccine-H gradually diminishes, and a greater health benefit can be achieved if more effort is exerted on achieving a wider reach. As the efficacy of vaccine-H increases, a lower level of IAR is achieved as more people are likely to become fully protected against the disease upon the vaccination (Figure S3). However, the rate of the reduction in IAR decreases in the efficacy for vaccine-H, and a relatively large reduction in IAR rather be achieved via increasing the reach ratio (Figure S4). If a decision-maker has the means of increasing the reach ratio by either reducing the resource requirements of vaccine-H relative to vaccine-L or increasing its capabilities to cover more individuals with vaccine-H given the resource requirements, this is always beneficial as it increases the number of vaccinated people with a limited amount of resources.

Increasing the reach ratio, however, may imply different levels of complexity depending on how it is defined in a given context. For example, low-income countries generally suffer from supply chain challenges (manufacturing, distribution, and storage) of vaccines against various diseases [29-32]. Many low- and, even, middle-income countries have faced supply chain challenges even more during the COVID-19 pandemic since mRNA vaccines, which are more effective and became available sooner than the other vaccine types, similar to the vaccine-H in our model, are produced based on a new vaccine technology and require colder temperatures than influenza vaccines [33, 34]. On the other hand, the potential solutions to these challenges, such as sharing vaccine technology with low- and middle-income countries and establishing an advanced cold-chain infrastructure, and, therefore, increasing the reach ratio, could be costly and time-consuming [35]. Such scenarios would correspond to the low values of the reach ratios in our models, where a decision-maker may not have much capability to change those ratios but treat as given. In such a case, the decision-maker can still minimize IAR by allocating more of its available resources to the lower efficacy vaccine given its potential wider reach, rather than putting a significant effort into trying to incrementally increase the reach ratio and allocate resources to vaccine-H (Figure 3). This also implies that if some individuals must be vaccinated early (e.g., frontline workers) but vaccine-H is too resource-intensive, the decision-maker should procure the smallest amount of vaccine-H that is enough to cover those individuals, which still results in a smaller IAR than investing the entire resources into vaccine-H and/or putting effort into increasing the reach ratio slightly.

Similar to the case of minimizing the IAR, our results identified the thresholds of the reach ratio, below which allocating the resources entirely to vaccine-L minimizes the peak percentage. However, this threshold is a lot smaller than the threshold that minimizes IAR (Table 2 and Figure S1). The implementation of effective vaccines as early as possible reduces the number of susceptible populations who could have had infectious contacts if the vaccines were not available. This leads to a slower rate of infections and a “flattened” curve of the pandemic [36]. When minimizing the peak percentage, policymakers often focus on the health benefits in the short-term, hoping the intervention prevents a large-scale infection within a small time of period rather than the overall infection levels throughout the course of the disease. If vaccine-H is moderately resource intensive so that allocating resources to vaccine-H minimizes the peak percentage but not the IAR, policymakers must make the resource allocation decision carefully according to the goal of the vaccination program. If the goal is to lower the peak percentage such that it would not exceed the local healthcare capacity but at the same time to reduce the infection levels over the long term, they may choose to allocate some resources to both vaccine types.

**Table 2:**
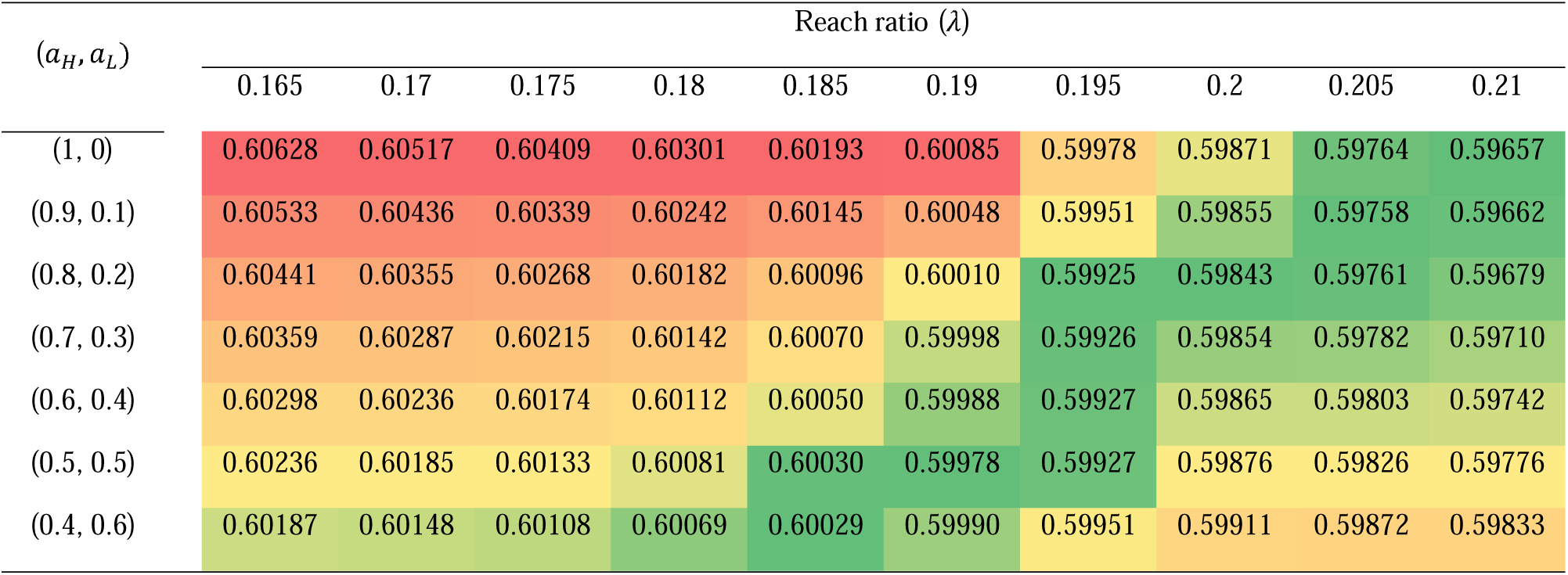
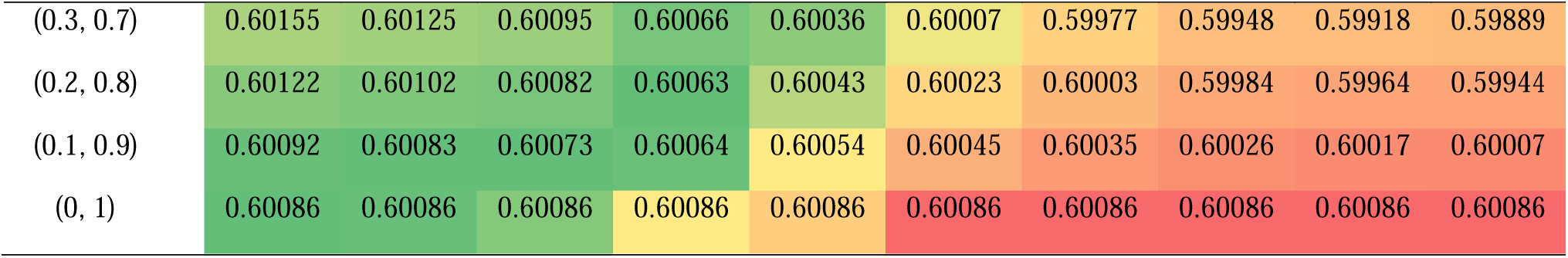
Peak percentage under different resource allocation decisions and reach ratios

| $(a_H, a_L)$ | Reach ratio ($\lambda$) | | | | | | | | | |
| --- | --- | --- | --- | --- | --- | --- | --- | --- | --- | --- |
|  | 0.165 | 0.17 | 0.175 | 0.18 | 0.185 | 0.19 | 0.195 | 0.2 | 0.205 | 0.21 |
| (1, 0) | 0.60628 | 0.60517 | 0.60409 | 0.60301 | 0.60193 | 0.60085 | 0.59978 | 0.59871 | 0.59764 | 0.59657 |
| (0.9, 0.1) | 0.60533 | 0.60436 | 0.60339 | 0.60242 | 0.60145 | 0.60048 | 0.59951 | 0.59855 | 0.59758 | 0.59662 |
| (0.8, 0.2) | 0.60441 | 0.60355 | 0.60268 | 0.60182 | 0.60096 | 0.60010 | 0.59925 | 0.59843 | 0.59761 | 0.59679 |
| (0.7, 0.3) | 0.60359 | 0.60287 | 0.60215 | 0.60142 | 0.60070 | 0.59998 | 0.59926 | 0.59854 | 0.59782 | 0.59710 |
| (0.6, 0.4) | 0.60298 | 0.60236 | 0.60174 | 0.60112 | 0.60050 | 0.59988 | 0.59927 | 0.59865 | 0.59803 | 0.59742 |
| (0.5, 0.5) | 0.60236 | 0.60185 | 0.60133 | 0.60081 | 0.60030 | 0.59978 | 0.59927 | 0.59876 | 0.59826 | 0.59776 |
| (0.4, 0.6) | 0.60187 | 0.60148 | 0.60108 | 0.60069 | 0.60029 | 0.59990 | 0.59951 | 0.59911 | 0.59872 | 0.59833 |
| (0.3, 0.7) | 0.60155 | 0.60125 | 0.60095 | 0.60066 | 0.60036 | 0.60007 | 0.59977 | 0.59948 | 0.59918 | 0.59889 |
| (0.2, 0.8) | 0.60122 | 0.60102 | 0.60082 | 0.60063 | 0.60043 | 0.60023 | 0.60003 | 0.59984 | 0.59964 | 0.59944 |
| (0.1, 0.9) | 0.60092 | 0.60083 | 0.60073 | 0.60064 | 0.60054 | 0.60045 | 0.60035 | 0.60026 | 0.60017 | 0.60007 |
| (0, 1) | 0.60086 | 0.60086 | 0.60086 | 0.60086 | 0.60086 | 0.60086 | 0.60086 | 0.60086 | 0.60086 | 0.60086 |

We acknowledge some limitations of this study. In our model, we used a simple compartment model to evaluate different strategies of resource allocation between different vaccine types without confounding the model with the effects of other interventions. However, the model can be extended to reflect disease transmission under such settings. In addition, our extended SIR-D model does not fully capture the potential trajectory of an infectious disease over its lifetime. There could be additional stages (compartments), such as *presymptomatic infected* individuals who are exposed to the virus but do not develop symptoms yet or *diagnosed/undiagnosed* individuals who are infected, get tested, and isolate themselves upon their decisions. Another model extension could be temporarily separating individuals who strictly follow non-pharmaceutical interventions from the susceptible population. Individuals who decide to stop conforming to the interventions may re-enter the susceptible populations during a pandemic, as considered in [37]. Some other extensions include a phased rollout of vaccines, instead of an immediate deployment as in our model, and different number of doses each vaccine type requires, etc.

Overall, our results suggest that allocating limited resources towards a vaccine with high efficacy that becomes available earlier than a vaccine with lower efficacy may not always result in increased benefits of a vaccine upon its implementation, especially if the latter can be distributed more widely. In fact, this may result in a significant deterioration in the infection attack rates if the high-efficacy vaccine is highly resource intensive, relative to the low-efficacy one, such that only a few people can be vaccinated each day. Therefore, identifying the resource intensity for each vaccine type as a function of their efficacy levels, timelines, and disease characteristics, is critical for resource allocation decisions, as there is a clear threshold for which vaccine type should be favored, and a significant improvement in health outcomes can be achieved. Manufacturing an mRNA-based vaccine is based on a new vaccine development technology and disseminating the vaccine have been challenging due to its stringent supply-chain requirements, especially in resource-limited countries. Improving the global access to life-saving vaccines by not only building a suitable infrastructure for effective distribution and storage of mRNA-based vaccines but also considering the tradeoffs and synergies between efficacy and reach is critical. We hope that this study can provide guidance to decision-makers in their resource planning for different vaccine types to better prepare for future pandemics.

## Data Availability

No data is used or referred to in the manuscript.

## ACKNOWLEDGEMENT

This research has been supported in part by the following Georgia Tech benefactors: William W. George, Andrea Laliberte, Claudia L. and J. Paul Raines, and Richard E. “Rick” and Charlene Zalesky. This research has also been supported in part by Cooperative Agreement number NU38OT000297 from The Centers for Disease Control and Prevention (CDC) and CSTE and does not necessarily represent the views of CDC and CSTE.

## ADDITIONAL INFORMATION

Dr. İnci Yildirim reported being a member of the mRNA-1273 Study Group. Dr. Yildirim has received funding to her institution to conduct clinical research from BioFire, MedImmune, Regeneron, PaxVax, Pfizer, GSK, Merck, Novavax, Sanofi-Pasteur, and Micron. Dr. Pinar Keskinocak received funding to her institution from Merck to conduct non-clinical research. The funders played no role in the study design, data collection, analysis, interpretation, or in writing the manuscript.

## AUTHOR CONTRIBUTIONS

D.K., P.K., P.P., and I.Y. conceived the model and contributed to the writing of the manuscript. D.K. contributed to the production of the figures and the tables.

## SUPPLEMENTARY MATERIALS

### A. Compartmental Model

We extended the susceptible-infected-recovered-deceased (SIR-D) compartmental model to capture the dynamics of COVID-19 transmission, isolation of symptomatic infected individuals, and implementation of two types of vaccines. The list below summarizes the compartments we included in the model:

1. Susceptible
  a. *S*: Susceptible
  b. *S*_*H*_: Population who has received vaccine-H but remains susceptible
  c. *S*_*L*_: Population who has received vaccine-L but remains susceptible
2. Infected
  a. *I*^*S*^: Symptomatic Infected
  b. *I*^*A*^: Asymptomatic Infected
  c. *Q*: Quarantined – infected individuals that do not transmit disease to susceptible populations
3. Vaccinated (Immunized)
  a. *V*_*H*_: Vaccinated with vaccine-H
  b. *V*_*L*_: Vaccinated with vaccine-L
4. Recovered
  a. *R*: Recovered
5. Deceased
  a. *D*: Deceased

Table S1 below summarizes the epidemiological and vaccine parameters, used in the extended SIR-D model:

**Table S1:** Epidemiological and vaccine parameters

| **Epidemiological Parameters** | Value<br>(main scenario) | Description |
| --- | --- | --- |
| $\beta^S$ | 0.21 | Rate of disease transmission from symptomatic-infected individuals |
| $\beta^A$ | 0.1575 | Rate of disease transmission from asymptomatic-infected individuals |
| $\gamma^S$ | 1/16 | Rate of recovery for symptomatic-infected individuals |
| $\gamma^A$ | 1/8 | Rate of recovery for asymptomatic-infected individuals |
| $\gamma^Q$ | 1/4 | Rate of recovery for quarantined individuals |
| $\phi$ | 1/12 | Rate of isolation by symptomatic-infected individuals |
| $\mu$ | 0.0015 | Rate of death among symptomatic-infected individuals |
| $p^S$ | 70% | Percentage of symptomatic infection |
| **Vaccine Parameters** | Value<br>(main scenario) | Description |
| $e_H$ | 90% | Efficacy for vaccine-H |
| $e_L$ | 70% | Efficacy for vaccine-L |
| $t_H$ | Day 1 | Time that vaccine-H becomes available |
| $t_L$ | Day 22 | Time that vaccine-L becomes available |
| $K$ | 1,000,000 | Daily resource capacity |
| $\lambda$ | [.005, .995; .005] | Reach ratio |
| $a_H$ | [0, 1; 0.1] | Decision of resource allocation to vaccine-H |
| $a_L$ | [0, 1; 0.1] | Decision of resource allocation to vaccine-L |
| $v_H$ | $a_H \lambda K / S$ | Percentage of susceptible individuals who receive vaccine-H |
| $v_L$ | $a_L K / S$ | Percentage of susceptible individuals who receive vaccine-L |

We used the R package *deSolve* to construct the simulation that utilizes the Euler method to solve the ordinary differential equations below with a step size of 1 day.

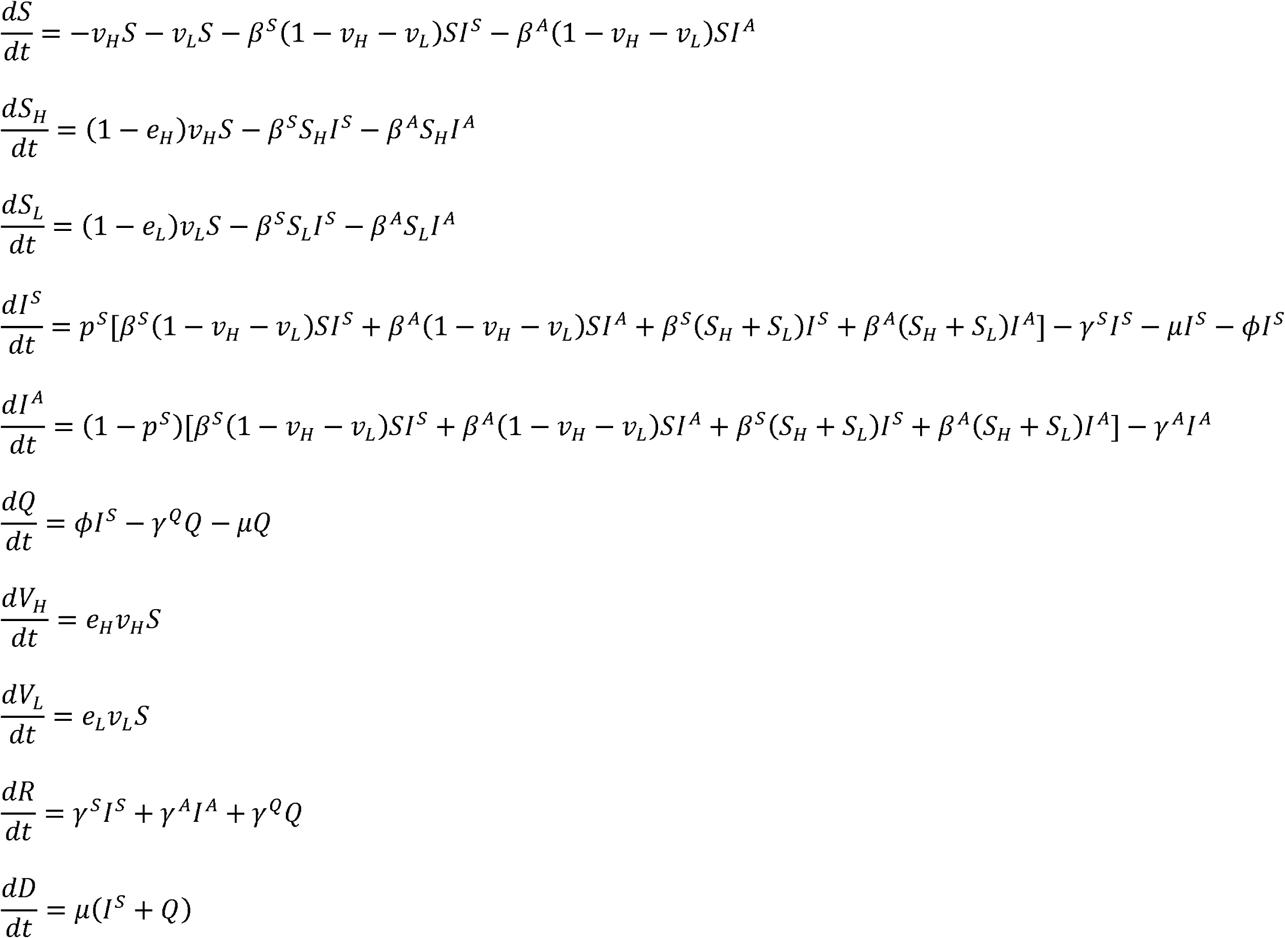

### B. Main Scenario – Full Results of the Simulation

Figure S1 shows the full results of the simulation under different reach ratios () and resource allocation decisions (when the main outcome of interest is either the infection attack rate (IAR) or the peak percentage.

**Figure S1:**
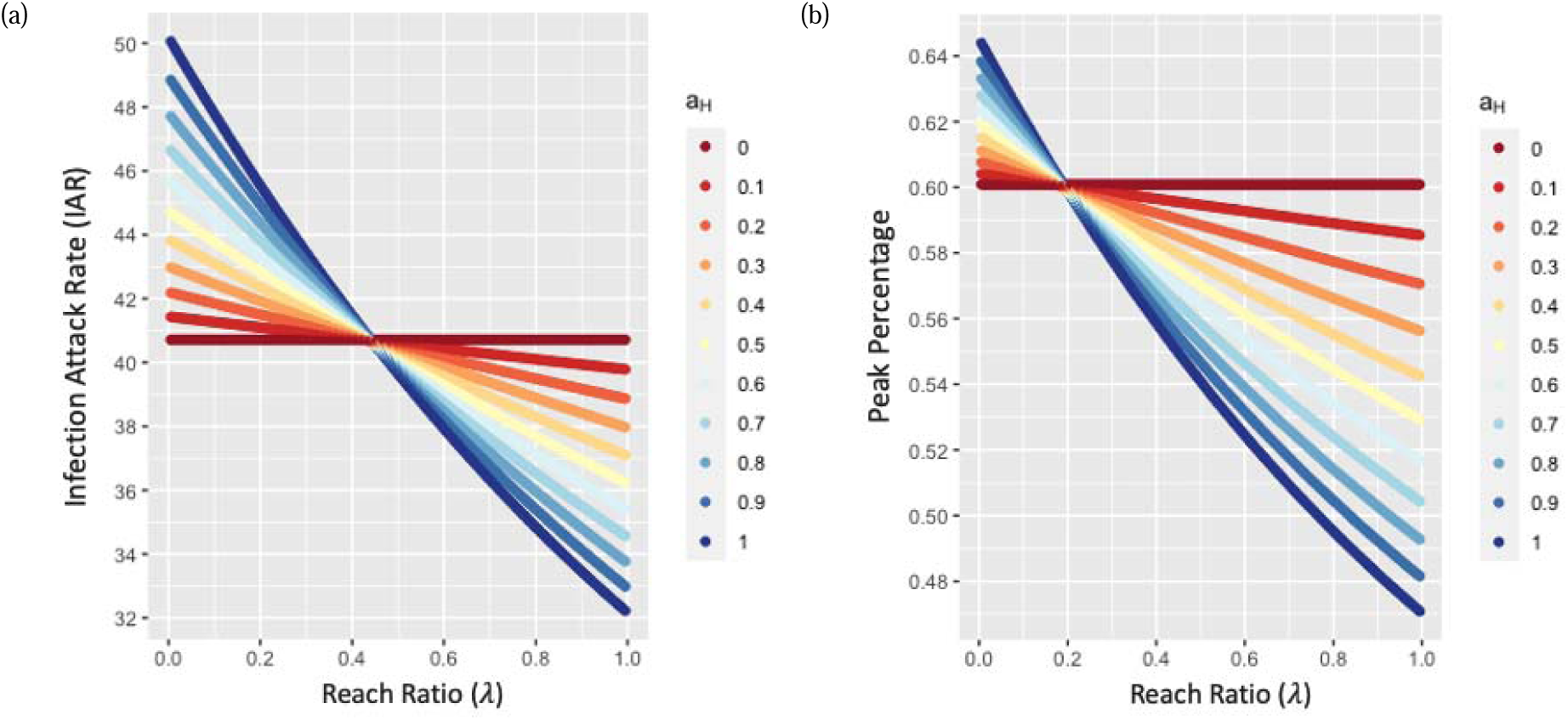
Main scenario simulation results when the objective is to minimize (a) infection attack rate (%) and (b) peak percentage (%)

### C. Sensitivity Analyses

We conducted extensive sensitivity analyses. A total of 21 additional scenarios were tested.

### a. The impact of the infectivity of a disease (*R*_0_)

Table S2 shows the IAR under different reach ratios and resource allocation decisions with a specified reproduction number (*R*_0_). It also reports the thresholds of the reach ratio, at or below which vaccine-L is favored entirely (*a*_*L*_ = 1) when minimizing IAR 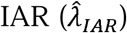 or peak percentage 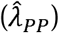, and the thresholds of the reach ratio, at or above which vaccine-H is favored entirely (*a*_*H*_ = 1) when minimizing IAR 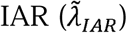 or peak percentage 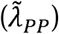, under different efficacies for vaccine-H. Similar to the main scenario, we observe that 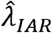 generally decreases as *R*_0_ increases. However, if vaccine-L becomes available after the peak day, we observe a lower 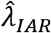. For example, under the scenario where vaccine-L becomes available on day 22, when *R*_0_ = 2.0, the peak day ranges between day 10 – 16, depending on the reach ratio and the resource allocation decision, and 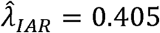 0.405. When *R*_0_ = 2.75, the peak day ranges between day 31 – 36 and 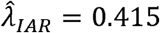.

**Table S2:** Infection Attack Rate (%) under different infectivity of a disease

| $R_0 = 2.0$ ($\beta^S = 0.169118, \beta^A = 0.1268385$) | | | | | | | | |
| --- | --- | --- | --- | --- | --- | --- | --- | --- |
| $(a_H, a_L)$ | Reach ratio ($\lambda$) | | | | | | | |
|  | 0.405 | 0.41 | 0.415 | 0.42 | 0.425 | 0.43 | 0.435 | 0.44 |
| (1, 0) | 20.635 | 20.587 | 20.539 | 20.492 | 20.445 | 20.399 | 20.353 | 20.307 |
| (0.9, 0.1) | 20.600 | 20.558 | 20.516 | 20.475 | 20.434 | 20.393 | 20.352 | 20.311 |
| (0.8, 0.2) | 20.571 | 20.534 | 20.498 | 20.461 | 20.426 | 20.390 | 20.354 | 20.319 |
| (0.7, 0.3) | 20.545 | 20.514 | 20.483 | 20.452 | 20.421 | 20.390 | 20.360 | 20.329 |
| (0.6, 0.4) | 20.524 | 20.498 | 20.472 | 20.446 | 20.419 | 20.394 | 20.368 | 20.342 |
| (0.5, 0.5) | 20.507 | 20.485 | 20.464 | 20.442 | 20.421 | 20.400 | 20.379 | 20.357 |
| (0.4, 0.6) | 20.493 | 20.476 | 20.459 | 20.442 | 20.425 | 20.408 | 20.392 | 20.375 |
| (0.3, 0.7) | 20.482 | 20.469 | 20.457 | 20.444 | 20.432 | 20.420 | 20.407 | 20.395 |
| (0.2, 0.8) | 20.474 | 20.466 | 20.458 | 20.449 | 20.441 | 20.433 | 20.425 | 20.417 |
| (0.1, 0.9) | 20.469 | 20.465 | 20.461 | 20.457 | 20.453 | 20.449 | 20.445 | 20.441 |
| (0, 1) | 20.466 | 20.466 | 20.466 | 20.466 | 20.466 | 20.466 | 20.466 | 20.466 |

| $(a_H, a_L)$ | Reach ratio ($\lambda$) | | | | | | |
| --- | --- | --- | --- | --- | --- | --- | --- |
|  | 0.425 | 0.43 | 0.435 | 0.44 | 0.445 | 0.45 | 0.455 |
| (1, 0) | 30.675 | 30.596 | 30.517 | 30.439 | 30.361 | 30.283 | 30.206 |
| (0.9, 0.1) | 30.622 | 30.552 | 30.482 | 30.413 | 30.344 | 30.276 | 30.207 |
| (0.8, 0.2) | 30.576 | 30.515 | 30.454 | 30.394 | 30.333 | 30.273 | 30.213 |
| (0.7, 0.3) | 30.537 | 30.484 | 30.432 | 30.380 | 30.328 | 30.276 | 30.224 |
| (0.6, 0.4) | 30.503 | 30.459 | 30.415 | 30.371 | 30.327 | 30.283 | 30.239 |
| (0.5, 0.5) | 30.476 | 30.439 | 30.403 | 30.367 | 30.331 | 30.295 | 30.259 |
| (0.4, 0.6) | 30.453 | 30.424 | 30.396 | 30.367 | 30.339 | 30.310 | 30.282 |
| (0.3, 0.7) | 30.436 | 30.414 | 30.393 | 30.372 | 30.351 | 30.330 | 30.309 |
| (0.2, 0.8) | 30.423 | 30.409 | 30.395 | 30.381 | 30.367 | 30.353 | 30.339 |
| (0.1, 0.9) | 30.414 | 30.408 | 30.401 | 30.394 | 30.387 | 30.380 | 30.373 |
| (0, 1) | 30.410 | 30.410 | 30.410 | 30.410 | 30.410 | 30.410 | 30.410 |

| $(a_H, a_L)$ | Reach ratio ($\lambda$) | | | | | |
| --- | --- | --- | --- | --- | --- | --- |
|  | 0.415 | 0.42 | 0.425 | 0.43 | 0.435 | 0.44 |
| (1, 0) | 51.735 | 51.641 | 51.548 | 51.455 | 51.361 | 51.268 |
| (0.9, 0.1) | 51.687 | 51.603 | 51.520 | 51.437 | 51.354 | 51.271 |
| (0.8, 0.2) | 51.644 | 51.571 | 51.498 | 51.424 | 51.351 | 51.278 |
| (0.7, 0.3) | 51.608 | 51.545 | 51.481 | 51.418 | 51.354 | 51.291 |
| (0.6, 0.4) | 51.578 | 51.524 | 51.470 | 51.416 | 51.363 | 51.309 |
| (0.5, 0.5) | 51.554 | 51.509 | 51.464 | 51.420 | 51.375 | 51.331 |
| (0.4, 0.6) | 51.534 | 51.499 | 51.463 | 51.428 | 51.393 | 51.358 |
| (0.3, 0.7) | 51.520 | 51.494 | 51.467 | 51.441 | 51.415 | 51.389 |
| (0.2, 0.8) | 51.510 | 51.493 | 51.476 | 51.458 | 51.441 | 51.424 |
| (0.1, 0.9) | 51.505 | 51.497 | 51.488 | 51.480 | 51.471 | 51.462 |
| (0, 1) | 51.505 | 51.505 | 51.505 | 51.505 | 51.505 | 51.505 |

**$R_0 = 3.0$ ( $\beta^S = 0.253677$ , $\beta^A = 19025775$ )**
| $(a_H, a_L)$ | Reach ratio ($\lambda$) | | | | | | |
| --- | --- | --- | --- | --- | --- | --- | --- |
|  | 0.395 | 0.4 | 0.405 | 0.41 | 0.415 | 0.42 | 0.425 |
| (1, 0) | 60.213 | 60.125 | 60.037 | 59.949 | 59.861 | 59.773 | 59.685 |
| (0.9, 0.1) | 60.165 | 60.087 | 60.008 | 59.929 | 59.851 | 59.772 | 59.694 |
| (0.8, 0.2) | 60.123 | 60.053 | 59.984 | 59.915 | 59.845 | 59.776 | 59.707 |
| (0.7, 0.3) | 60.086 | 60.026 | 59.965 | 59.905 | 59.845 | 59.785 | 59.725 |
| (0.6, 0.4) | 60.054 | 60.003 | 59.952 | 59.900 | 59.849 | 59.798 | 59.747 |
| (0.5, 0.5) | 60.027 | 59.985 | 59.942 | 59.900 | 59.858 | 59.816 | 59.773 |
| (0.4, 0.6) | 60.005 | 59.971 | 59.938 | 59.904 | 59.871 | 59.837 | 59.804 |
| (0.3, 0.7) | 59.988 | 59.963 | 59.938 | 59.913 | 59.888 | 59.863 | 59.838 |
| (0.2, 0.8) | 59.975 | 59.958 | 59.942 | 59.925 | 59.908 | 59.892 | 59.875 |
| (0.1, 0.9) | 59.966 | 59.958 | 59.950 | 59.941 | 59.933 | 59.925 | 59.917 |
| (0, 1) | 59.961 | 59.961 | 59.961 | 59.961 | 59.961 | 59.961 | 59.961 |

The peak day is day 30 – 33, depending on the reach ratio ( $\lambda$ ) and the resource allocation decision ( $a_H$ and $a_L$ ). When $a_L = 1$ , the peak day is day 31.

### b. The impact of the time that vaccine-L becomes available ()

In Figure S2, each contour plot shows the infection attack rate (IAR) under different reach ratios, (x-axis), and resource allocation to vaccine-H, (y-axis) with a specified time that vaccine-L becomes available (). In each contour plot, there is a reach ratio, at which the shape of the contour line changes from convex-increasing to convex-decreasing, which is denoted as. To minimize the IAR, (i.e.,) if and otherwise. As increases, the moves leftward, indicating the case that more resource-intensive vaccine-H receives the entire allocation as the time that vaccine-L becomes available gets delayed.

**Figure S2:**
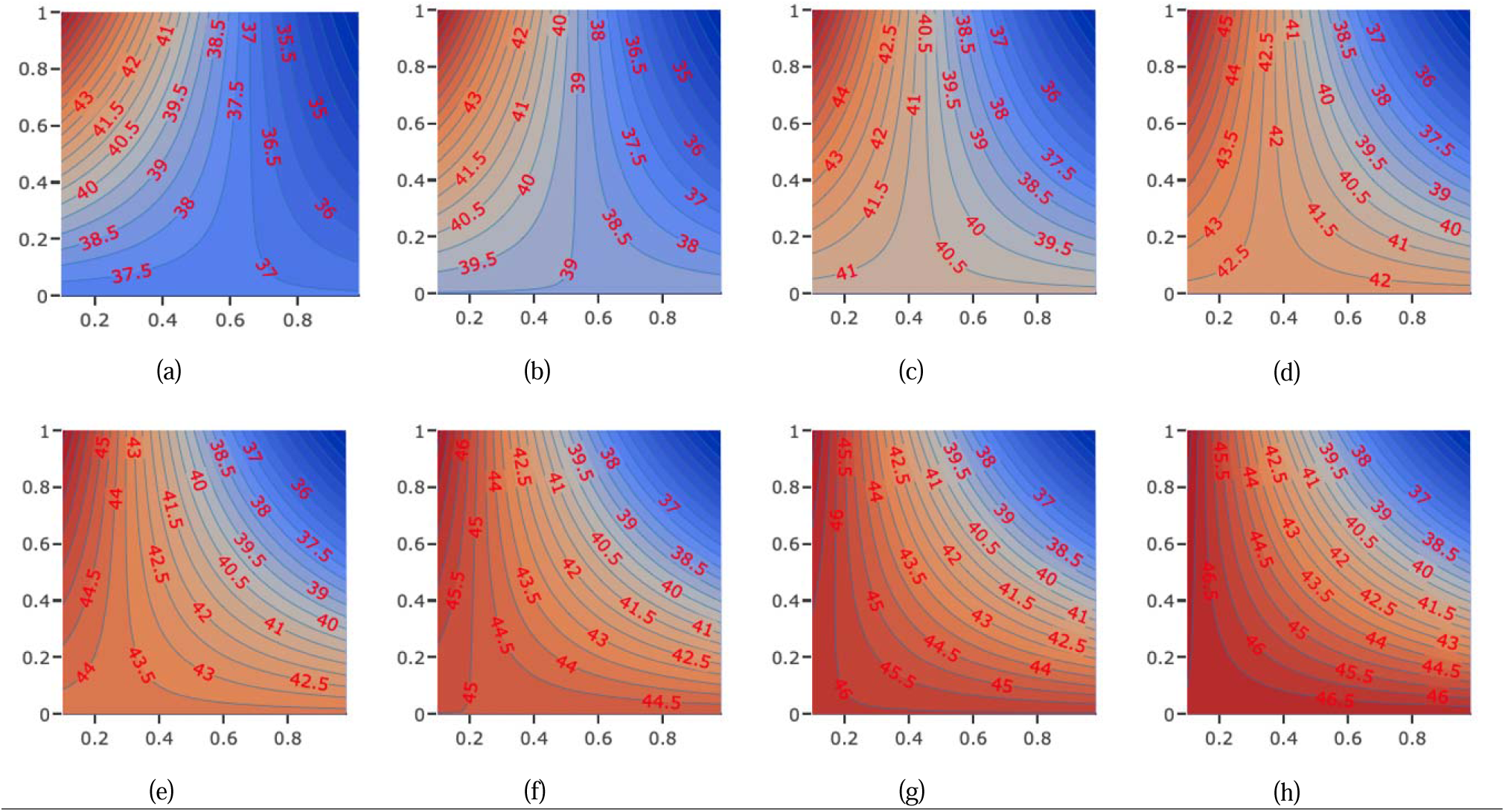
Contour plots of infection attack rate (x-axis: reach ratio, ; y-axis: resource allocation to vaccine-H,) under different times that vaccine-L becomes available.

Table S3 shows 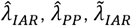 and 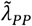 under different times that vaccine-L becomes available. When the reach ratio is between 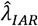 and 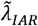, there exists an optimal combination of resource allocation to both vaccine types (i.e., 0 < *a*_*H*_, *a*_*L*_ < 1) that minimizes the IAR. Similarly, when the reach ratio is between 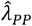 and 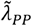, there exists an optimal combination of resource allocation to both vaccine types (i.e., 0 < *a*_*H*_, *a*_*L*_ < 1) that minimizes the peak percentage. Note that when *t*_*L*_ > 43, the peak percentage when *a*_*L*_ = 1 does not change. This is because vaccine-L becomes available after the peak day, and, therefore, the peak percentage is the same as in the absence of any vaccine. When *t*_*L*_ = 36, 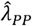 does not exist, which indicates that vaccine-H receives partial or entire allocation even if it is extremely resource-intensive. When *t*_*L*_ > 43, 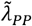 does not exist; this indicates that vaccine-H receives the entire resource allocation no matter how resource-intensive it is.

**Table S3:** Identification of reach ratio thresholds under different times that vaccine-L becomes available

| Vaccine-L<br>Time of Availability ($t_L$) | IAR<br>$a_L = 1$ | Peak % (Day)<br>$a_L = 1$ | $\hat{\lambda}_{IAR}$ | IAR<br>$a_H = 1^1$ | $\tilde{\lambda}_{IAR}$ | IAR<br>$a_H = 1^2$ | $\hat{\lambda}_{PP}$ | Peak % (Day)<br>$a_H = 1^3$ | $\tilde{\lambda}_{PP}$ | Peak % (Day)<br>$a_H = 1^4$ |
| --- | --- | --- | --- | --- | --- | --- | --- | --- | --- | --- |
| 1<sup>5</sup> | 35.070 | 0.49653 (31) | 0.780 | 35.130 | 0.785 | 35.058 | 0.790 | 0.49661 (31) | 0.795 | 0.49592 (31) |
| 8 | 37.081 | 0.53495 (32) | 0.645 | 37.195 | 0.655 | 37.035 | 0.530 | 0.53495 (33) | 0.540 | 0.53416 (33) |
| 15 | 38.968 | 0.57021 (33) | 0.530 | 39.116 | 0.550 | 38.771 | 0.320 | 0.57396 (36) | 0.350 | 0.56809 (35) |
| 22 | 40.716 | 0.60086 (34) | 0.425 | 41.009 | 0.455 | 40.454 | 0.165 | 0.60628 (38) | 0.210 | 0.59657 (37) |
| 29 | 42.311 | 0.62535 (36) | 0.335 | 42.743 | 0.375 | 41.959 | 0.070 | 0.62811 (39) | 0.095 | 0.62225 (38) |
| 36 | 43.740 | 0.64289 (37) | 0.270 | 44.062 | 0.310 | 43.244 | - | - | 0.020 | 0.64029 (39) |
| 43 | 44.994 | 0.64521 (39) | 0.200 | 45.548 | 0.250 | 44.480 | - | - | - | - |
| 50 | 46.070 | 0.64521 (39) | 0.155 | 46.539 | 0.200 | 45.548 | - | - | - | - |
| 57 | 46.970 | 0.64521 (39) | 0.115 | 47.445 | 0.160 | 46.428 | - | - | - | - |

<sup>1</sup> IAR under $\lambda = \hat{\lambda}_{IAR}$ and $a_H = 1$
<sup>2</sup> IAR under $\lambda = \tilde{\lambda}_{IAR}$ and $a_H = 1$
<sup>3</sup> Peak percentage under $\lambda = \hat{\lambda}_{PP}$ and $a_H = 1$
<sup>4</sup> Peak percentage under $\lambda = \tilde{\lambda}_{PP}$ and $a_H = 1$
<sup>5</sup> This scenario assumes that vaccine-L and vaccine-H become available on the same day. Unlike any other scenarios, the optimal allocation decision in this scenario can be inferred without an epidemiological model. A decision-maker can minimize the IAR by choosing a resource allocation such that the number of individuals who are vaccinated and fully protected against the disease on each day is maximized (i.e., $(a_H, a_L)$ s.t. $e_H a_H \lambda K + e_L a_L K$ is maximized).

### c. The impact of the efficacy levels of vaccine-H ()

Figure S3 shows the change in infection attack rate (IAR) with respect to the efficacy level of vaccine-H when the reach ratio is fixed at 0.43. IAR when is labeled on the figure. For a linear increase in the efficacy level of vaccine-H, the reduction in IAR slowly decreases for all. The result stays robust under different reach ratios.

**Figure S3:**
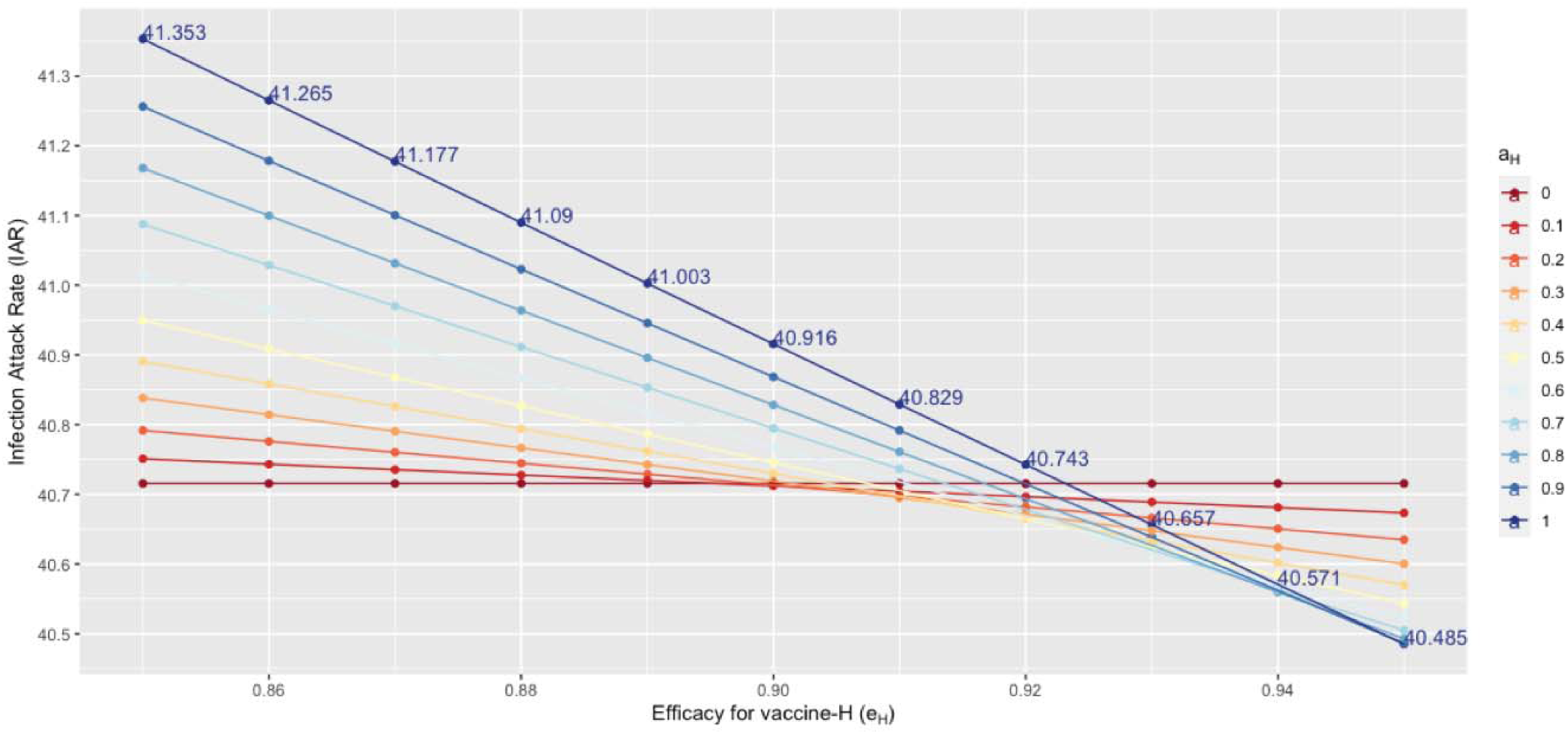
Infection attack rate (%) with respect to the efficacy for vaccine-H when

Figure S4 shows the IAR under different efficacy levels of vaccine-H () and reach ratio). Under a fixed, IAR decreases as increases. The rate of reduction in IAR, however, decreases as increases. Under a fixed, a higher leads to a greater reduction in IAR.

**Figure S4:**
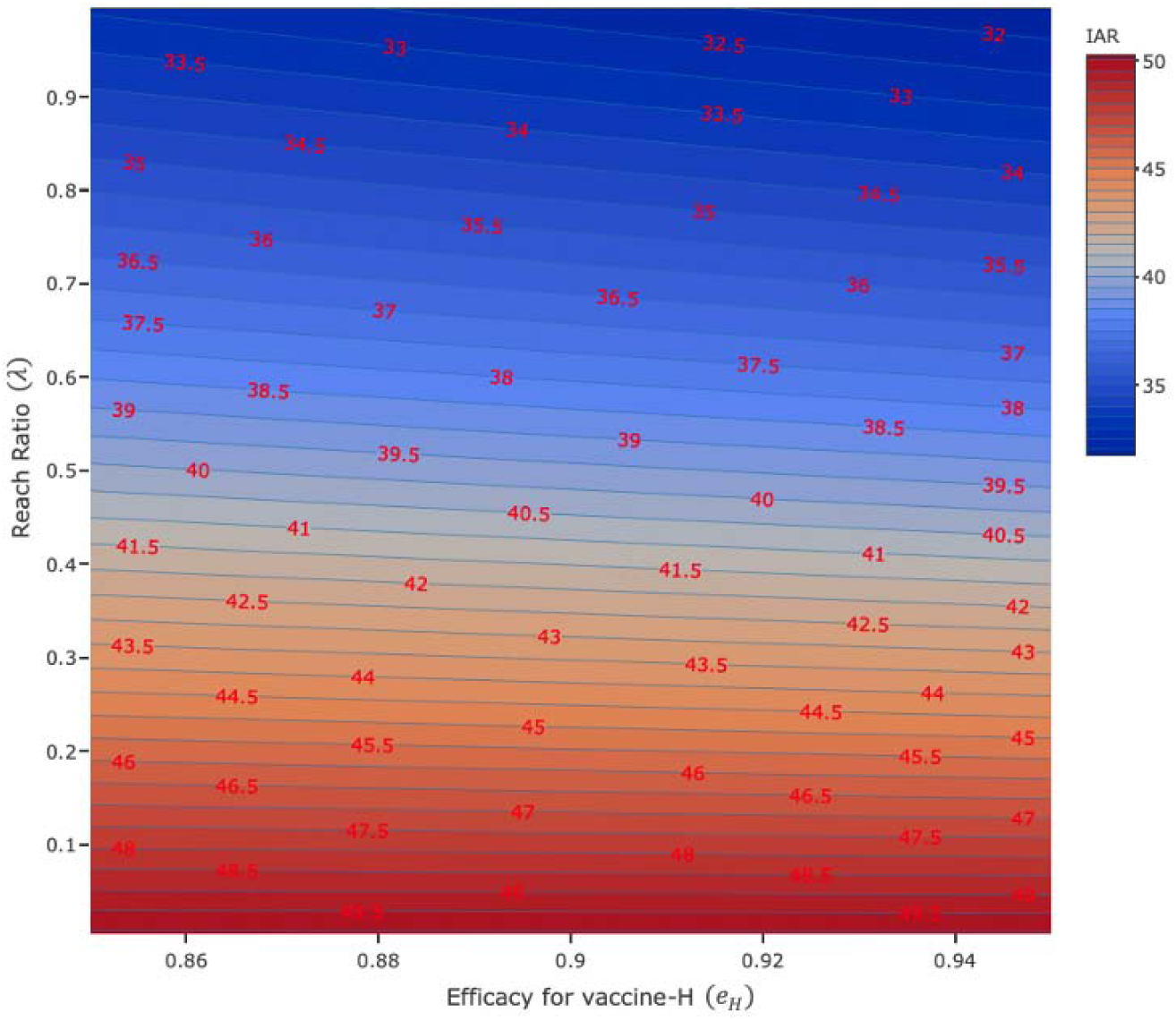
Infection attack rate (%) with respect to the efficacy for vaccine-H

Table S4 shows 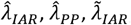, and 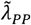 under different efficacies for vaccine-H. Both vaccines are allocated resources if the reach ratio is between 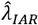 and 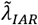 (or between 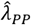 and 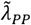). When a_L_ = 1, the level of infection attack rate is 40.716% and the peak percentage is 0.60086% on day 34, regardless of the level of efficacy of vaccine-H. As the efficacy level increases, all the reach ratio thresholds decrease, and the IAR and peak percentage when a_H_ = 1 decrease.

**Table S4:** Identification of reach ratio thresholds under different efficacies for vaccine-H

| Vaccine-H<br>Efficacy (%) | Vaccine-L |  |  |  | Vaccine-H |  |  |  |
| --- | --- | --- | --- | --- | --- | --- | --- | --- |
| | $\hat{\lambda}_{IAR}$ | IAR<br>when $a_H = 1$ | $\hat{\lambda}_{PP}$ | Peak % (Day)<br>when $a_H = 1$ | $\tilde{\lambda}_{IAR}$ | IAR<br>when $a_H = 1$ | $\tilde{\lambda}_{PP}$ | Peak % (Day)<br>when $a_H = 1$ |
| 85 | 0.450 | 40.998 | 0.175 | 0.60612 (38) | 0.480 | 40.472 | 0.220 | 0.59692 (37) |
| 86 | 0.445 | 40.996 | 0.175 | 0.60570 (38) | 0.475 | 40.465 | 0.220 | 0.59642 (37) |
| 87 | 0.440 | 40.996 | 0.170 | 0.60637 (38)** | 0.47 | 40.459 | 0.215 | 0.59696 (37) ** |
| 88 | 0.435 | 40.998* | 0.170 | 0.60596 (38) | 0.465 | 40.455 | 0.215 | 0.59647 (37) |
| 89 | 0.43 | 41.003* | 0.165 | 0.60667 (38) ** | 0.46 | 40.454 | 0.210 | 0.59704 (37) ** |
| 90 | 0.425 | 41.009* | 0.165 | 0.60628 (38) | 0.455 | 40.454 | 0.210 | 0.59657 (37) |
| 91 | 0.42 | 41.017* | 0.165 | 0.60588 (38) | 0.45 | 40.456* | 0.210 | 0.59609 (37) |
| 92 | 0.415 | 41.028* | 0.160 | 0.60664 (38) ** | 0.445 | 40.461* | 0.205 | 0.59671 (37) ** |
| 93 | 0.41 | 41.040* | 0.160 | 0.60626 (38) | 0.44 | 40.467* | 0.205 | 0.59624 (37) |
| 94 | 0.41 | 40.957 | 0.160 | 0.60587 (38) | 0.435 | 40.475* | 0.200 | 0.59689 (37) ** |
| 95 | 0.405 | 40.972* | 0.155 | 0.60668 (38) ** | 0.43 | 40.485* | 0.200 | 0.59644 (37) |

\*: The reported IAR level in the table is higher than the IAR under a lower vaccine-H efficacy ( $e_H$ ). This is because, as $e_H$ increases, the threshold of the reach ratio ( $\hat{\lambda}_{IAR}$ and $\tilde{\lambda}_{IAR}$ ), decreases. Under the same reach ratio $\lambda$ , the IAR decreases as $e_H$ increases (e.g., under $\lambda = 0.44$ and $e_H = 88\%$ , the IAR when $a_H = 1$ is 40.907%, which is lower than the IAR of 40.996% under $\lambda = 0.44$ and $e_H = 87\%$ ).
\*\*: The reported peak percentage in the table is higher than the peak percentage under a lower vaccine-H efficacy ( $e_H$ ). This is because, as $e_H$ increases, the threshold of the reach ratio ( $\hat{\lambda}_{PP}$ and $\tilde{\lambda}_{PP}$ ), decreases. Under the same reach ratio $\lambda$ , the peak percentage decreases as $e_H$ increases.

### D. Alternative Scenarios

We considered alternative scenarios, changing the settings of the main scenario: when (i) vaccine-H became available later than vaccine-L and (ii) vaccine-H became available sooner than vaccine-L as in the main scenario but both vaccines were delayed.

### a. Vaccine-H becomes available later than vaccine-L

In this alternative scenario, keeping all other parameters the same as in the main scenario, we made vaccine-L became available on day 1 and varied the time that vaccine-H became available (later than vaccine-L). We also varied the reach ratio within the range of 0.005 and 3.00 with a discrete step size of 0.005 in this alternative scenario.

**Table S5:** Reach ratio thresholds under different times that vaccine-H becomes available

| Vaccine-H<br>Available Time | $\hat{\lambda}_{IAR}$ | IAR<br>$a_H = 1$ | $\hat{\lambda}_{PP}$ | Peak % (Day)<br>$a_H = 1$ | $\tilde{\lambda}_{IAR}$ | IAR<br>$a_H = 1$ | $\tilde{\lambda}_{PP}$ | Peak % (Day)<br>$a_H = 1$ |
| --- | --- | --- | --- | --- | --- | --- | --- | --- |
| 8 | 0.945 | 35.178 | 1.200 | 0.49923 (28) | 0.960 | 35.010 | 1.275 | 0.49381 (28) |
| 15 | 1.175 | 35.298 | 2.185 | 0.50814 (26) | 1.235 | 34.809 | > 3.0 | ---- |
| 22 | 1.490 | 35.612 | > 3.0 | ---- | 1.720 | 34.341 | > 3.0 | ---- |
| 29 | 1.945 | 36.083 | > 3.0 | ---- | 2.765 | 33.362 | > 3.0 | ---- |
| The peak percentage when the corresponding threshold exceeds 3.00 is omitted |  |  |  |  |  |  |  |  |

When a_L_ = 1 (i.e., *a*_*H*_ = 0), IAR is 35.070% and the peak percentage is 0.49653% on day 31. These results are not affected by the time that vaccine-H becomes available because vaccine-H receives no allocation.

### b. Both vaccines become available later than in the main scenario

i. Scenario-A
  - Initial Condition:
    Both vaccine-H and vaccine-L become available 14 days later than in the main scenario (i.e., day 15 and day 36, respectively).
    On day 15, the population size of each compartment is as follows:

    Cumulative number of cases is 10.42 % and the active number of cases is 2.797 % of the total population.

|  |  |
| --- | --- |
| $S$ | 89.199% |
| $I^S$ | 1.873% |
| $I^A$ | 0.924% |
| $Q$ | 0.544% |
| $R$ | 7.323% |
| $D$ | 0.137% |
| $V_H, V_L, S_H, S_L$ | 0% |

**Table S6:** Infection attack rate (IAR) in percentage under different reach ratios and resource allocation decisions when vaccine-H and vaccine-L become available 14 days later than in the main scenario
ii. **Scenario-B**
  - Initial Condition:
    Both vaccine-H and vaccine-L become available 23 days later than in the main scenario (i.e., day 24 and day 45, respectively).
    On day 24, the population size of each compartment is as follows:

| $(a_H, a_L)$ | Reach ratio ($\lambda$) | | | | | | |
| --- | --- | --- | --- | --- | --- | --- | --- |
|  | 0.375 | 0.38 | 0.385 | 0.39 | 0.395 | 0.40 | 0.405 |
| (1, 0) | 44.164 | 44.094 | 44.025 | 43.957 | 43.888 | 43.820 | 43.752 |
| (0.9, 0.1) | 44.117 | 44.056 | 43.995 | 43.934 | 43.873 | 43.812 | 43.752 |
| (0.8, 0.2) | 44.077 | 44.023 | 43.969 | 43.916 | 43.863 | 43.810 | 43.757 |
| (0.7, 0.3) | 44.042 | 43.996 | 43.949 | 43.903 | 43.857 | 43.811 | 43.766 |
| (0.6, 0.4) | 44.013 | 43.973 | 43.934 | 43.895 | 43.856 | 43.817 | 43.779 |
| (0.5, 0.5) | 43.988 | 43.956 | 43.924 | 43.892 | 43.860 | 43.827 | 43.796 |
| (0.4, 0.6) | 43.968 | 43.943 | 43.917 | 43.892 | 43.867 | 43.841 | 43.816 |
| (0.3, 0.7) | 43.953 | 43.934 | 43.915 | 43.896 | 43.877 | 43.859 | 43.840 |
| (0.2, 0.8) | 43.941 | 43.929 | 43.917 | 43.904 | 43.892 | 43.879 | 43.867 |
| (0.1, 0.9) | 43.934 | 43.928 | 43.922 | 43.916 | 43.909 | 43.903 | 43.897 |
| (0, 1) | 43.930 | 43.930 | 43.930 | 43.930 | 43.930 | 43.930 | 43.930 |

The peak day is day 17 – 23, depending on the reach ratio ( $\lambda$ ) and the resource allocation decision ( $a_H$ and $a_L$ ). When $a_L = 1$ , the peak day is day 23.

    Cumulative number of cases is 15.07% and the active number of cases is 3.586% of the total population.

|  |  |
| --- | --- |
| $S$ | 84.439% |
| $I^S$ | 2.403% |
| $I^A$ | 1.183% |
| $Q$ | 0.720% |
| $R$ | 11.081% |
| $D$ | 0.174% |
| $V_H, V_L, S_H, S_L$ | 0% |

**Table S7:**
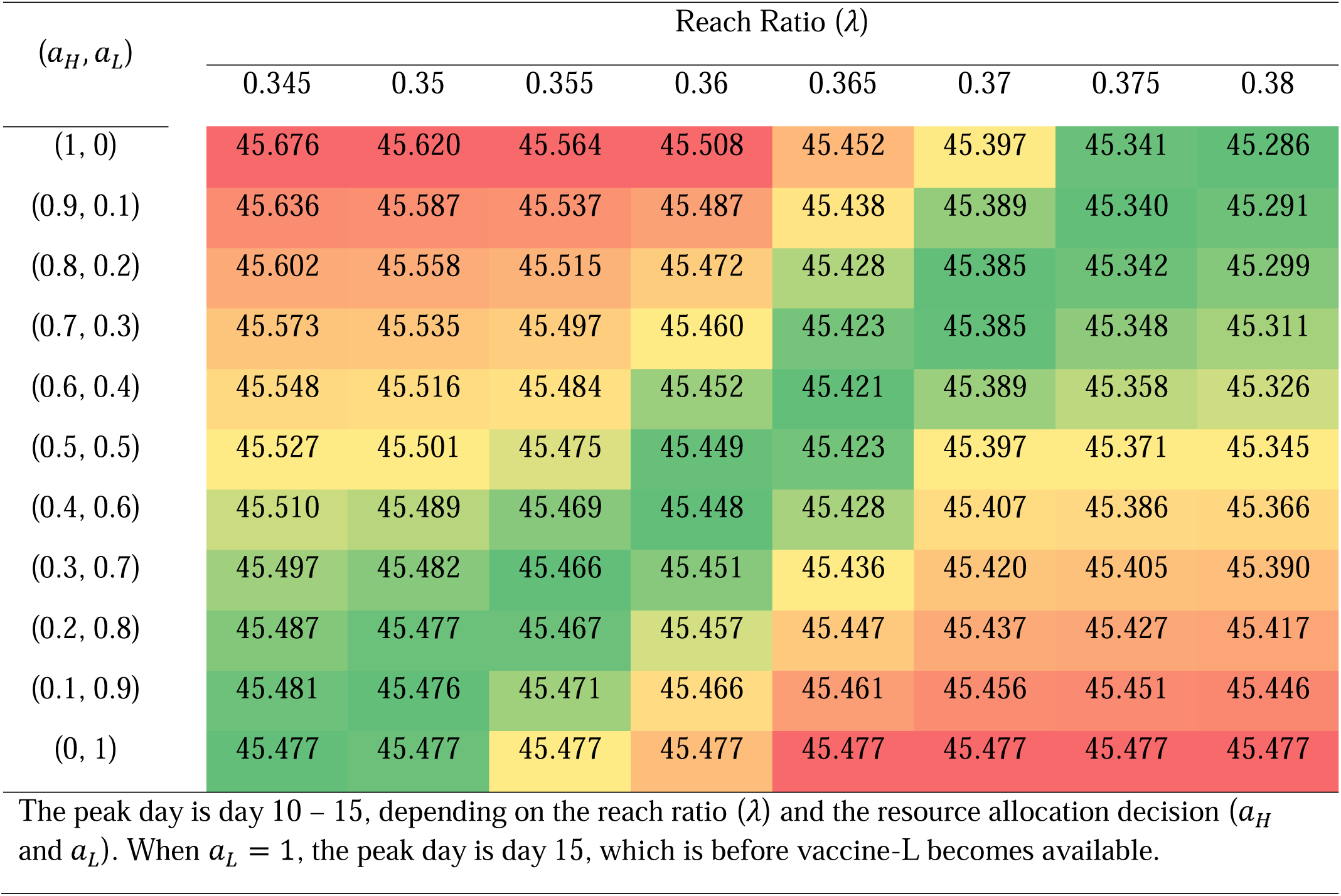
Infection attack rate (IAR) in percentage under different reach ratios and resource allocation decisions when vaccine-H and vaccine-L become available 23 days later than in the main scenario

| $(a_H, a_L)$ | Reach Ratio ($\lambda$) | | | | | | | |
| --- | --- | --- | --- | --- | --- | --- | --- | --- |
|  | 0.345 | 0.35 | 0.355 | 0.36 | 0.365 | 0.37 | 0.375 | 0.38 |
| (1, 0) | 45.676 | 45.620 | 45.564 | 45.508 | 45.452 | 45.397 | 45.341 | 45.286 |
| (0.9, 0.1) | 45.636 | 45.587 | 45.537 | 45.487 | 45.438 | 45.389 | 45.340 | 45.291 |
| (0.8, 0.2) | 45.602 | 45.558 | 45.515 | 45.472 | 45.428 | 45.385 | 45.342 | 45.299 |
| (0.7, 0.3) | 45.573 | 45.535 | 45.497 | 45.460 | 45.423 | 45.385 | 45.348 | 45.311 |
| (0.6, 0.4) | 45.548 | 45.516 | 45.484 | 45.452 | 45.421 | 45.389 | 45.358 | 45.326 |
| (0.5, 0.5) | 45.527 | 45.501 | 45.475 | 45.449 | 45.423 | 45.397 | 45.371 | 45.345 |
| (0.4, 0.6) | 45.510 | 45.489 | 45.469 | 45.448 | 45.428 | 45.407 | 45.386 | 45.366 |
| (0.3, 0.7) | 45.497 | 45.482 | 45.466 | 45.451 | 45.436 | 45.420 | 45.405 | 45.390 |
| (0.2, 0.8) | 45.487 | 45.477 | 45.467 | 45.457 | 45.447 | 45.437 | 45.427 | 45.417 |
| (0.1, 0.9) | 45.481 | 45.476 | 45.471 | 45.466 | 45.461 | 45.456 | 45.451 | 45.446 |
| (0, 1) | 45.477 | 45.477 | 45.477 | 45.477 | 45.477 | 45.477 | 45.477 | 45.477 |

The peak day is day 10 – 15, depending on the reach ratio ( $\lambda$ ) and the resource allocation decision ( $a_H$ and $a_L$ ). When $a_L = 1$ , the peak day is day 15, which is before vaccine-L becomes available.

